# Predictors of first-dose COVID-19 vaccine uptake in Benin: Evidence from a cross-sectional study based on a telephonic survey

**DOI:** 10.1101/2025.03.12.25323828

**Authors:** Elias Martinien Avahoundje, Christelle Boyi Hounsou, Kéfilath Bello, Armelle Akouavi Vigan, Christian M Agossou, Mena K. Agbodjavou, Ibrahima Gaye, Mouhamadou Faly Ba, Adama Faye, Valéry Ridde, Jean Paul Dossou

**Affiliations:** Centre de Recherche en Reproduction Humaine et en Démographie (CERRHUD), Cotonou, Benin; Institute of Health and Development (ISED), Cheikh Anta Diop University, Dakar, Senegal; Faculty of Medicine, Pharmacy and Odontology Cheikh Anta Diop University, Dakar, Senegal; Université Paris Cité, IRD, Inserm, Ceped, F-75006 Paris, France

**Keywords:** COVID-19, vaccine uptake, Benin, Determinants, Public health, National survey

## Abstract

**Introduction:** Vaccination against COVID-19 is an essential tool in the fight against the pandemic, but adherence to vaccination programmes is a major challenge, particularly in African countries. This study aimed to identify determinants of uptake of the first dose of COVID-19 vaccine using large-scale data collected from the Beninese population.

**Methods:** We conducted a cross-sectional and analytical study using a telephone survey between December 2021 and January 2022. The study covered Beninese aged 18 and over. Marginal quota sampling (n = 858) was used, with age, gender and department as quota variables. The questionnaire was inspired by the Theory of Planned Behaviour and the Health Belief Model. Binary logistic regression was used at the 5% significance level.

**Results:** According to this study, 56.9% of people had received at least one dose of the COVID-19 vaccine. The likelihood of getting the first dose is increased by having confidence in the vaccine (aOR= 2.009; CI= 1.414-2.853), finding the length of waiting time at vaccination centres acceptable (aOR= 1.601; CI= 1.128-2.273), living in the centre of Benin (aOR= 2. 398, IC= 1.461-3.935), not having heard or seen anything bad about the vaccine (aOR= 1.586 IC= 1.444-2.200), and having a high perceived benefit (aOR= 1.57; IC: 1.003-2.458). However, the main barriers to vaccination were never having been vaccinated as an adult (aOR= 0.463; CI: 0.333-0.643), being over 60 (aOR= 0.428; CI: 0.220-0.832), and perceiving the risk of vaccination as high (aOR= 0.669; CI: 0.481-0.931).

**Conclusions:** This study has provided results that can guide decision-makers in epidemic response programmes against COVID-19 and other similar pandemics Vaccination coverage is not negligible, but efforts are still needed, particularly in the south, among the elderly and people without previous vaccination experience. It is also important to improve confidence in the vaccine and to combat misinformation.

## Introduction

The COVID-19 pandemic severely impacted the global health landscape. Between November 2019 and February 2024, more than 774,631,444 positive cases associated with COVID-19, and 7,031,216 deaths were reported worldwide [1]. This change in the number of victims over time has been a major challenge, requiring nations to respond quickly and effectively. Considerable efforts have been made to make the first vaccines available one year after the pandemic’s start, in December 2020 [2]. The development of vaccines has been a major step forward in the fight against the virus. Recent studies have demonstrated the effectiveness of vaccines in preventing severe forms of COVID-19, including those associated with the variants of the COVID-19 virus (Alpha, Beta, Gamma and Delta) [3,4]. However, the success of vaccination campaigns depends not only on the availability of vaccines, but also, and above all, on the public’s willingness to be vaccinated.

Indeed, after the first vaccines were launched, reluctance towards all vaccines was noted among many people in several countries, hindering their widespread acceptance and adoption by the population [5]. Unfortunately, inadequate vaccination practices often lead to a resurgence of infectious diseases and an increase in mortality rates, as was the case with polio in Nigeria [6,7]. It has been shown that a large proportion of the population needs to be vaccinated to significantly reduce the spread of COVID-19. Some studies recommend a vaccination coverage rate of at least 75% [8]. Aware of the various challenges to the success of COVID-19 vaccination campaigns, several studies have identified the determining factors in vaccine intention [9,10]. However, it should be noted that the initial intention to vaccinate does not always translate into actual vaccination [11]. This is much more relevant in the COVID-19 pandemic, where the rapid evolution of the pandemic may lead to changes in the public’s perception or attitude to the virus [12,13]. Faced with these different situations, researchers set out to understand the factors that do or do not encourage people to be effectively vaccinated against COVID-19.

Thus, the literature reports various factors that can influence vaccination status against COVID-19. These include not only socio-demographic factors such as religion [14,15], ethnicity [16], age [17,18], gender [19], level of education [20] but also factors related to beliefs and behaviour towards the vaccine. These factors include perceived risk[21], attitudes and knowledge about the vaccine [17], and subjective norms [14]. Unvaccinated people are more likely to have unfavourable attitudes towards vaccination against COVID-19, concerns about side effects and long-term repercussions, and a lack of confidence in the safety and efficacy of the vaccine[16,17,22–28]. A study carried out in 15 countries around the world revealed that people who trust the government to manage the pandemic, believe in the health authorities and recognise the seriousness of the virus are more inclined to complete their vaccine doses than others [29]. Fear of contracting a COVID-19 infection [30] is also a factor in favour of vaccination. Confidence in decision-makers also encourages vaccination in Kenya [20].

In Benin, after campaigns were launched on 29 March 2021, public uptake did not meet expectations. By 27 February 2022, 2,897,882 doses of vaccine had been administered, representing a coverage of about 27% (at least one dose), far below the 60% expected at that time [31]. These results may be partly attributable to resistance in the Beninese population after the first vaccines became available [32]. In the literature on determinants of actual vaccination against COVID-19, most studies have been carried out in North-American, European and Asian countries. Very few studies exist in sub-Saharan Africa, particularly in French-speaking countries [28,33]. Also, many studies have focused on specific groups rather than the general population [27,34,35]. To our knowledge, to date only one study has been carried out on this subject in few communes (Abomey-Calavi, Cotonou, Djougou, Porto-Novo) in Benin but others have defined the dependent variable for the analysis as a combination of vaccination status and vaccination intention [36].

This study aims to fill this gap by identifying the determinants of receiving at least one dose of COVID-19 using data collected on a large scale from the Beninese population.

## Materiel and method

### Setting

Benin is a West African country with an estimated population of 12,314,650 in 2022 [37]. In 2021, Benin ranked 166^th^ out of 191 countries, with a Human Development Index of 0.525 [38]. In 2022, only 34.4% of households had access to the internet and 84.9% had a mobile or fixed telephone [39].

Benin recorded its first case of COVID-19 in March 2020. In February 2022, 2,836 cases of COVID-19 had been reported, and 163 people had died [1]. The vaccination campaign began about a year after the first case was recorded (March 2021). The campaign, which is part of the national vaccination programme, was supported by the COVAX (COVID-19 Vaccines Global Access) initiative, which facilitated the introduction of the AstraZeneca/Oxford and Sinovac vaccines, the establishment of vaccination sites and the development of vaccine administration protocols [40]. Pfizer, Moderna and Johnson Johnson vaccines became available a few weeks after the start of the campaign.

### Study design and population

We conducted a quantitative cross-sectional study repeated (once). Our study population consisted of Beninese men and women aged 18 and over. We have conducted two telephone surveys. The first data collection took place from 30 March to 15 May 2021 and collected complete information from 865 people. The second data collection (from 19 December 2021 to 20 January 2022) was carried out on the same people who had participated in the first phase and who had agreed to be contacted for a second data collection. The analyses in this publication are based on data collected in phase 2.

### Sampling and data collection

The sampling methodology used in the first phase and the technology employed are documented in a previous publication [10,41]. In summary, this cross-sectional survey was conducted using a marginal quota sampling method. This approach is particularly effective in emergency situations, especially when the sample size is less than 3,000 [42]. Quota variables were age, gender and department of origin. Quotas for each subgroup are proportional to Benin’s population distribution. Individuals were contacted using telephone numbers randomly generated by a specialized computer system. Eligibility, as well as the completion of the various quotas, was determined using information from the predefined quota variables. Of the 865 people who participated in the first survey, 552 completed the phase 2. Inaccessible participants were contacted 5 (five) times over 5 days before exclusion.

New automatically generated telephone numbers were contacted in order achieve a sufficiently large sample size and fill all quotas. A total of 306 additional participants completed the questionnaire, giving a sample size of 858 for the second survey. As mentioned above, the analyses in this study are based on the 858 respondents who took part in phase 2. It should be noted that not all quotas were met in all sub-groups. We therefore performed a calibration using the raking ratio method, in order to adjust the data to the population structure of Benin projected for 2022[43].

### Data collection techniques

We used a digital questionnaire on a tablet using the Open Data Kit (ODK) programme. The questionnaire took around 45 minutes to administer. Five interviewers were recruited and trained to conduct the interviews. The interviews were conducted in French, and in several local languages: Fon, Yoruba, Adja, Dindi, Bariba and Mina. A questionnaire pre-test was carried out before the launch of each of the two surveys, and two supervisors were recruited to monitor and control the quality of the data collected in real time.

### Theoretical framework

The information gathered in this study in relation to vaccination is inspired by two theoretical models often used to explain individuals’ health behaviour : the Theory of Planned Behaviour (TPB) [44] and the Health Belief Model (HBM) [45]. TPB maintains that health behaviour is determined by the individual’s intention to adopt it. This intention is a function of the individual’s attitude (the extent to which the individual is favourable or unfavourable to the behaviour), subjective norms (social influences and the perception that individuals have of what those close to them might think of their behaviour) and perceived behavioural control (the ease or difficulty the individual perceives in performing the behaviour). The HBM maintains that health behaviour is determined by the perceived severity of a particular health problem, perceived sensitivity to the problem, effectiveness, perceived benefits of preventive action and perceived obstacles to action (perceived risks, perceived difficulties, etc.).

### Dependent variable

The dependent variable in our study is vaccination status against COVID-19. It was captured only in phase 2 by the question: ‘Have you received a vaccine against COVID-19?’ This question was not asked in phase 1 because the survey began just 1 day after the launch of the vaccination campaign in Benin. It has two modalities. 1 ‘Get at least one dose’ and 0 ‘Not get at least on dose’’. In this study, we consider vaccinated persons to have received at least the first dose of any COVID-19 vaccine.

### Explanatory variables

We collected information on socio-demographic characteristics (age, sex, level of education, marital status, department of origin, household assets). The questionnaires also collected information on vaccine attitudes, subjective norms, behavioural control, perceived benefits, perceived risks, perceived efficacy, emotions, behavioural control, and confidence in the COVID-19 vaccine and in the provider who would administer a COVID-19 vaccine. Each factor was measured by a set of sub-questions (Supplementary material). For each dimension, Cronbach’s alpha, an internal consistency parameter, was calculated to test the reliability of the measurement instruments. Most of the values obtained were above 0.6 (Table 1), which indicates that the instrument is highly reliable [46]. This also shows that the items used to measure each component are linked. We also collected data on previous experience with vaccination, respondents’ opinions on appropriate opening hours and the acceptable or unacceptable length of the queues at the vaccination centres. Whether or not they had heard anything bad about the COVID-19 vaccine was also recorded. All the questions were closed and treated as categorical variables. Most of the answers to the questions on vaccination were given on a five-point Likert scale indicating their degree of agreement from 1 (strongly disagree) to 5 (strongly agree).

**Table 1:** Demographic and health characteristics of the study population.

| Variables | n(%) |
| --- | --- |
| **Region** |  |
| North | 286(33.3) |
| Center | 126(14.7) |
| South | 446(52.0) |
| **Gender** |  |
| Male | 414(48.3) |
| Female | 444(51.7) |
| **Age Group** |  |
| Under 25 years | 316(36.8) |
| 25 - 59 years | 481(56.1) |
| 60 years and above | 61(7.1) |
| **Marital Status\*** |  |
| In union | 531(62.1) |
| Not in union | 325(37.9) |
| **Education Level** |  |
| Less educated | 360(42.0) |
| More educated | 498(58.0) |
| **Social Well-being** |  |
| Low | 595(69.3) |
| Medium | 155(18.1) |
| High | 108(12.6) |
| **Previous vaccine experience in adulthood** |  |
| Yes | 537(62.6) |
| No | 321(37.4) |
| **Vaccination status** |  |
| Not get at least on dose | 370(43.1) |
| Get at least one dose | 488(56.9) |
| **Total** | **858(100)** |

\*Two missing values for these variables, bringing the total to 856 in these cases.

### Data analysis

As part of the analyses, we used confirmatory factor analysis (CFA) to create latent variables that capture the different dimensions of the TPB and HBM. With CFA, when the latent variable is used as an explanatory variable in a regression, the estimates obtained from them are very slightly affected by measurement error bias [47]. The latent variables obtained represent composite indicators that record scores. The higher the score, the stronger the individual’s perception of the measured factor. For example, individuals with higher scores on the ‘perceived risk’ dimension have a higher perception of the risk associated with vaccination. We then recoded each latent variable into two modalities: 0 ‘low’ and 1 ‘high’. This is low when the score on the composite variable is lower than the average score, and high when the opposite is true. The CFA produces indicators that make it possible to assess the adjustment quality at the level of each latent variable created. We have the comparative fit index (CFI) > 0.90, the root mean square error of approximation (RMSEA) < 0.05 and the standardised mean residual (SRMR) < 0.08 [48]. Table A.1 shows that the coefficients obtained for each variable are satisfactory.

We carried out a descriptive and explanatory analysis. In descriptive analysis, we presented the socio-demographic variables to get an overview of the study population. Since all our variables were categorical, a chi2 test was performed to test whether there was an association between the dependent variable and the various explanatory variables. In the explanatory analysis, a binary logistic regression model was estimated to identify the factors determining vaccination status. The explanatory variables introduced into the regression model had a p-value of less than 0.25 in the bivariate analysis. The model produces adjusted odds ratios that allow us to assess the associations between the explanatory variables and COVID-19 vaccination status. The significance level used was 5%. We tested for multicolinearity between the different variables. The test gave a value of VIF=1.23, which is synonymous with an absence of multicolinearity because this value is less than 2.5 [49]. The Hosmer & Lemeshow test was also used to measure the goodness of fit of the model. We ranked the determining factors from the logistic regression using the relative contribution of each variable on the basis of the variation in the likelihood ratio chi-square statistic. All analyses were performed with STATA 16 software.

## Ethical considerations

This study was approved by the Local Ethics Committee for Biomedical Research of the University of Parakou in Benin under number: 0308/CLERB-UP/P/SP/R/SA. We also obtained the statistical visa of the National Institute of Statistics and Demography of Benin under number: 21/2020/MPD/INSAE/DCSFR. Oral informed consent was also obtained from each respondent to the questionnaire during both surveys.

## Results

### Description of the study population

The results in Table 1 show that of the 858 people who participated in our study, most respondents were from the South of Benin (52%). Most were aged between 25 and 59 (56.1%), better educated (58.0%), in union (62.1%) and had a low standard of living (69.3%). Many people had once received a vaccine other than COVID-19 in adulthood (69.3%). We note that 56.9% of our study population had received at least one dose of COVID-19 vaccine at the time of data collection.

### Two-way association between socio-demographic characteristics and vaccination status

Table 2 shows a significant association between vaccination status and region and previous vaccine receipt in adulthood at the 5% threshold. The vaccination rate is higher in the centre and north than in the south. There was no significant association between age group, gender, level of education, marital status, or socio-economic well-being at the 5% threshold. In addition the following variables: Subjective norms, attitude, behavioural control (ability), perceived effectiveness, perceived benefit, perceived risk, positive emotion, acceptable queue length, adequate opening hours, having seen or heard something negative about COVID-19 vaccines, trusting the provider who would administer a COVID-19 vaccine, having confidence in the new COVID-19 vaccine if it were available now are significantly associated with vaccination status.

**Table 2:** Bivariate analysis between study variables and COVID-19 vaccination status in 2022.

|  | Not get at least on dose<br>n(%) | Get at least one<br>dose<br>n(%) | N | P-value |
| --- | --- | --- | --- | --- |
| **Region** |  |  |  | **0.002** |
| North | 121(42.4) | 165(57.6) | 286 |  |
| Center | 35(27.7) | 91(72.3) | 126 |  |
| South | 214(47.9) | 233(52.1) | 446 |  |
| **Gender** |  |  |  | 0.704 |
| Male | 182(43.9) | 232(56.1) | 414 |  |
| Female | 188(42.4) | 256(57.6) | 444 |  |
| **Age Group** |  |  |  | 0.246 |
| Under 25 years | 132(41.8) | 184(58.2) | 316 |  |
| 25 - 59 years | 205(42.5) | 277(57.5) | 481 |  |
| 60 years and above | 33(55.2) | 27(44.8) | 60 |  |
| **Marital Status\*** |  |  |  | **0.089** |
| In union | 216(40.7) | 315(59.3) | 531 |  |

|  | Not get at least on dose | Get at least one dose |  |  |
| --- | --- | --- | --- | --- |
|  | n(%) | n(%) | N | P-value |
| Not in union | 154(47.4) | 171(52.6) | 325 |  |
| Total | 370(43.2) | 486(56.8) | 856 |  |
| **Education Level** |  |  |  | 0.390 |
| Less educated | 148(41.2) | 212(58.8) | 360 |  |
| More educated | 222(44.5) | 276(55.5) | 498 |  |
| **Social-economique Well-being** |  |  |  | 0.598 |
| Low | 254(42.8) | 340(57.2) | 595 |  |
| Medium | 64(41.1) | 92(58.9) | 155 |  |
| High | 52(48) | 56(52) | 108 |  |
| **Previous vaccine experience in adulthood** |  |  |  | <0.001 |
| Yes | 188(35) | 349(65) | 537 |  |
| No | 182(56.7) | 139(43.3) | 321 |  |
| **Level of subjective norms** |  |  |  | <0.001 |
| Low | 146(60.7) | 95(39.3) | 240 |  |
| High | 224(36.3) | 394(63.7) | 618 |  |
| **Attitudes towards the vaccine** |  |  |  | <0.001 |
| Unfavourable | 112(70.6) | 47(29.4) | 159 |  |
| Favourable | 258(36.9) | 441(63.1) | 699 |  |
| **Behavioural control: ability** |  |  |  | <0.001 |
| Low | 156(58.4) | 111(41.6) | 267 |  |
| High | 214(36.3) | 377(63.7) | 591 |  |
| **Behavioural control: autonomy** |  |  |  | 0.394 |
| No | 28(37.7) | 47(62.3) | 75 |  |
| Yes | 342(43.7) | 441(56.3) | 783 |  |
| **Perceived effectiveness** |  |  |  | <0.001 |
| Low | 171(57.5) | 126(42.5) | 297 |  |
| High | 199(35.5) | 362(64.5) | 561 |  |
| **Perceived benefit** |  |  |  | <0.001 |
| Low | 145(68.9) | 66(31.1) | 211 |  |
| High | 225(34.7) | 423(65.3) | 647 |  |
| **Perceived risk** |  |  |  | <0.001 |
| Low | 148(32.3) | 311(67.7) | 459 |  |
| High | 222(55.6) | 177(44.4) | 399 |  |
| **Negative emotions** |  |  |  | 0.923 |
| Low | 108(42.8) | 145(57.2) | 253 |  |
| High | 262(43.2) | 343(56.8) | 605 |  |
| **Positive Emotion** |  |  |  | 0.060 |
| Low | 91(50.1) | 91(49.9) | 182 |  |
| High | 279(41.2) | 397(58.8) | 676 |  |
| **Acceptable queue length** |  |  |  | <0.001 |
| No | 181(57.2) | 135(42.8) | 316 |  |
| Yes | 190(34.9) | 353(65.1) | 543 |  |
| **Adequate opening hours** |  |  |  | <0.001 |
| No | 111(66.9) | 55(33.1) | 166 |  |

|  | Not get at least on dose | Get at least one dose | N | P-value |
| --- | --- | --- | --- | --- |
|  | n(%) | n(%) |  |  |
| Yes | 259(37.4) | 433(62.6) | 692 |  |
| **Seen or heard anything bad about COVID-19 vaccines** |  |  |  | **<0.001** |
| Yes | 237(49.3) | 244(50.7) | 482 |  |
| No | 133(35.3) | 244(64.7) | 376 |  |
| **Trust the provider who would give a COVID-19 vaccine** |  |  |  | **<0.001** |
| No | 119(70.2) | 51(29.8) | 170 |  |
| Yes | 251(36.5) | 437(63.5) | 688 |  |
| **Confidence in the new COVID-19 vaccine if it were available to you now** |  |  |  | **<0.001** |
| No | 239(61.3) | 151(38.7) | 390 |  |
| Yes | 131(28) | 337(72) | 468 |  |
| Total | 370(43.1) | 488(56.9) | 858 |  |

### Determinants of vaccination status

The information in Table 3 and Table 4 shows that the determining factors for vaccination status, in order of importance, are the fact of having already received a vaccine in adulthood, confidence in the vaccine, region, having heard something bad about the COVID-19 vaccine, satisfaction with the length of the queue at vaccination sites, age, perceived risk and perceived benefit. Respondents who had not been vaccinated as adults (aOR= 0.463; CI: 0.333-0.643), those aged 60 and over compared with those aged under 25 (aOR= 0.428; CI: 0.220-0.832) and those with a high perception of vaccine-related risk (aOR= 0.669; CI: 0.481-0.931) were less likely to receive at least one dose of one of the COVID-19 vaccines. In contrast, respondents who had confidence in the vaccine (aOR= 2.009; CI= 1.414-2.853), those who had a good assessment of the length of the waiting time at vaccination centres (aOR= 1.601; CI= 1.128-2.273), those living in the centre of Benin compared with those living in the south (aOR= 2. 398, IC= 1.461-3.935), those who had not seen or heard anything negative about the vaccine (aOR=1.586 IC= 1.444-2.200) and those with a high perceived benefit (aOR= 1.57; IC: 1.003-2.458) were more likely to receive at least one dose.

**Table 3:**
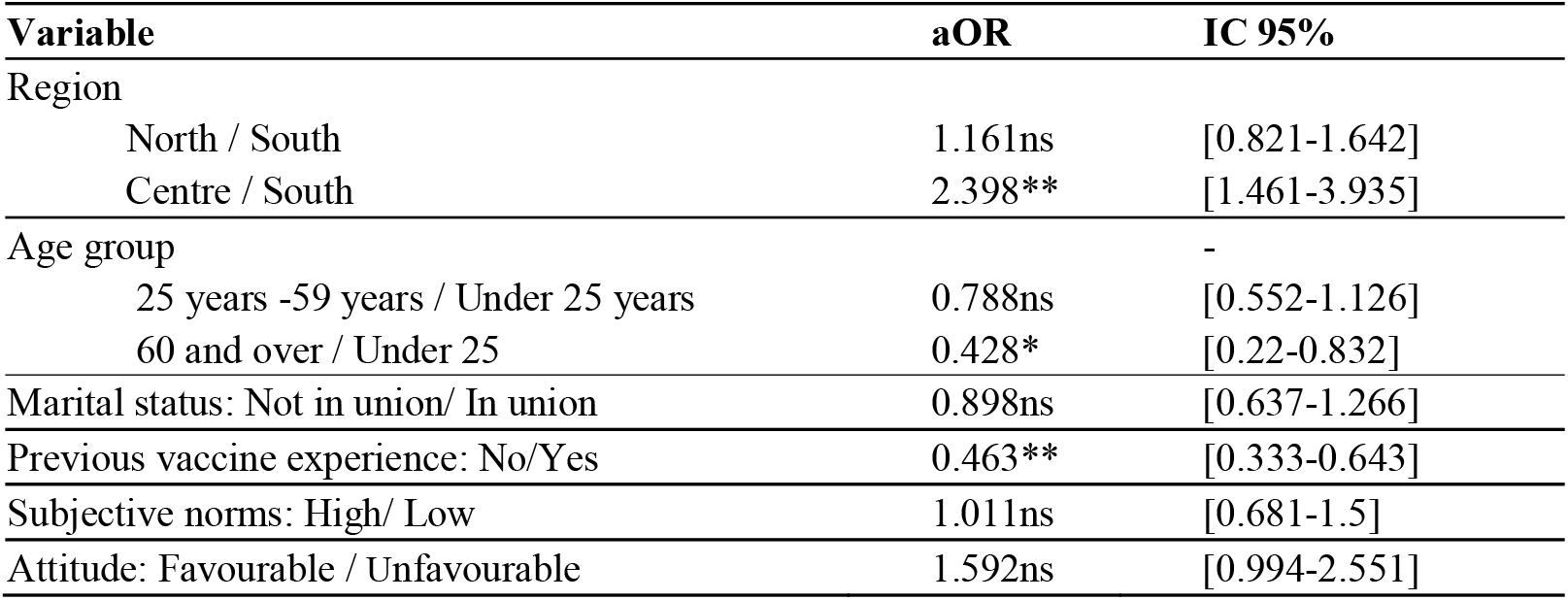
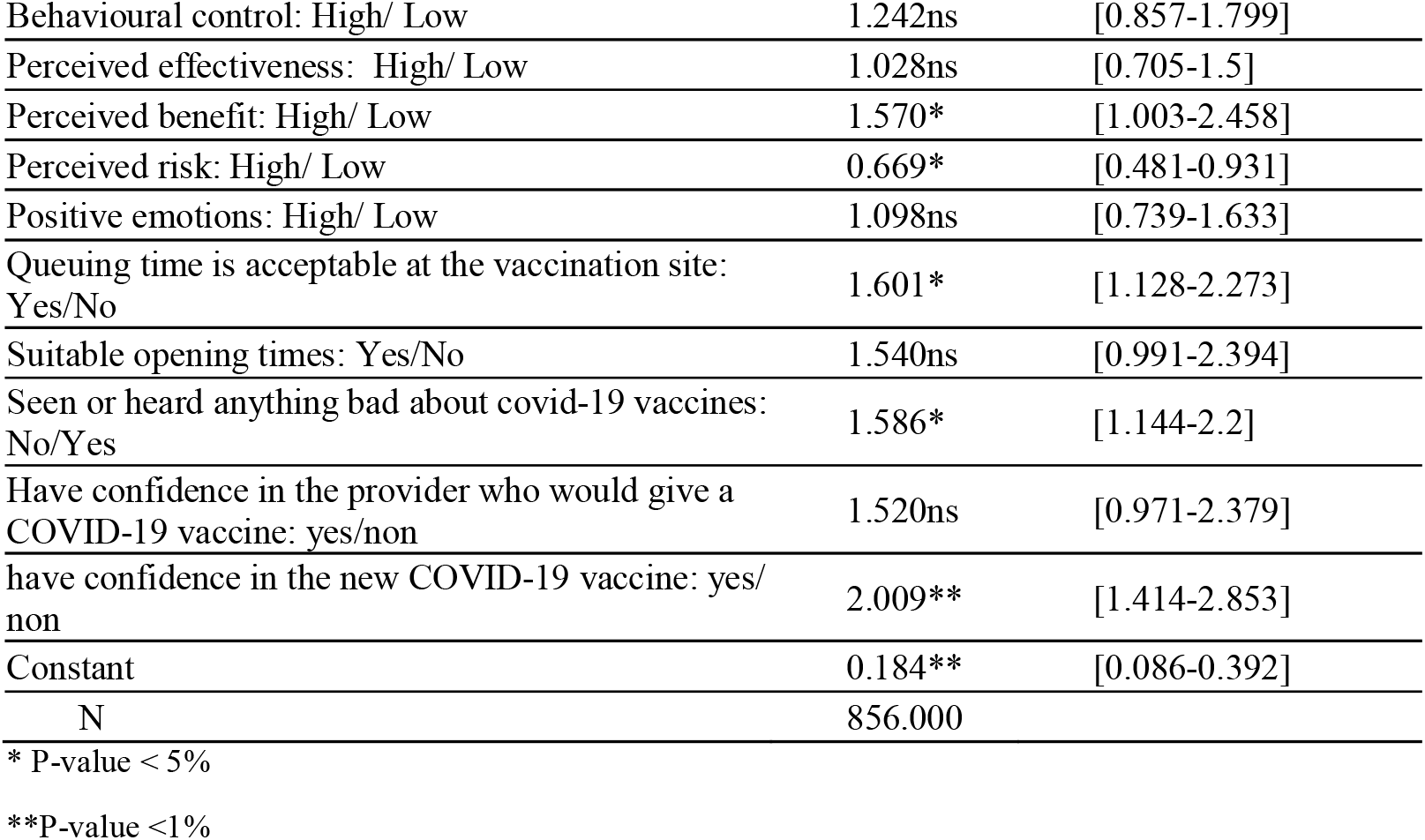
Logistic regression to identify determinants of uptake of first dose of COVID-19 vaccine, 2022.

| Variable | aOR | IC 95% |
| --- | --- | --- |
| Region |  |  |
| North / South | 1.161ns | [0.821-1.642] |
| Centre / South | 2.398** | [1.461-3.935] |
| Age group |  | - |
| 25 years -59 years / Under 25 years | 0.788ns | [0.552-1.126] |
| 60 and over / Under 25 | 0.428* | [0.22-0.832] |
| Marital status: Not in union/ In union | 0.898ns | [0.637-1.266] |
| Previous vaccine experience: No/Yes | 0.463** | [0.333-0.643] |
| Subjective norms: High/ Low | 1.011ns | [0.681-1.5] |
| Attitude: Favourable / Unfavourable | 1.592ns | [0.994-2.551] |
| Behavioural control: High/ Low | 1.242ns | [0.857-1.799] |
| Perceived effectiveness: High/ Low | 1.028ns | [0.705-1.5] |
| Perceived benefit: High/ Low | 1.570* | [1.003-2.458] |
| Perceived risk: High/ Low | 0.669* | [0.481-0.931] |
| Positive emotions: High/ Low | 1.098ns | [0.739-1.633] |
| Queuing time is acceptable at the vaccination site: Yes/No | 1.601* | [1.128-2.273] |
| Suitable opening times: Yes/No | 1.540ns | [0.991-2.394] |
| Seen or heard anything bad about covid-19 vaccines: No/Yes | 1.586* | [1.144-2.2] |
| Have confidence in the provider who would give a COVID-19 vaccine: yes/non | 1.520ns | [0.971-2.379] |
| have confidence in the new COVID-19 vaccine: yes/non | 2.009** | [1.414-2.853] |
| Constant | 0.184** | [0.086-0.392] |
| N | 856.000 |  |

\* P-value < 5%
\*\*P-value <1%

**Table 4:** Hierarchisation of determinants.

| Variables | LR Chi2 of model without the variable (a) | LR Chi2 of full model (b) | Relative contribution (c) | Rank |
| --- | --- | --- | --- | --- |
| Previous vaccine experience | 204.168 | 225.361 | 9.4% | 1 |
| Have confidence in the new COVID-19 vaccine | 210.313 | 225.361 | 6.7% | 2 |
| Region | 212.684 | 225.361 | 5.6% | 3 |
| Seen or heard anything bad about covid-19 vaccines | 217.647 | 225.361 | 3.4% | 4 |
| Queuing time is acceptable at the vaccination site | 218.48 | 225.361 | 3.1% | 5 |
| Age Group | 218.869 | 225.361 | 2.9% | 6 |
| Perceived risk | 219.726 | 225.361 | 2.5% | 7 |
| Perceived benefit | 221.482 | 225.361 | 1.7% | 8 |

c=(b-a)/b

## Discussion

Our study showed that 56.9% of the population studied had received at least one dose of one of the COVID-19 vaccines. This is slightly higher than the national rate of 27% reported by official government statistics during the study period. It should be noted that the present study focused only on Beninese aged 18 and over. Initially, vaccination was only available to Beninese aged 18 and over. However, the calculation of the official rate is based on the general population. Knowing that the African population has a fairly broad pyramid base (over 50% of Benin’s population is under 18 [43]), the official rate should logically be lower than that which would have been obtained if the population aged 18 and over had been considered.

Previous vaccination experience appears to be the primary determinant of vaccine status. Individuals who have already been vaccinated as adults are more likely to be vaccinated against COVID-19. A study carried out in a border community between the United States and Mexico showed that people who had received the influenza vaccine were more likely to be vaccinated against COVID-19 [50]. A systematic review of childhood influenza vaccine uptake came to similar conclusions [51]. It has also been shown in South Africa that previous influenza vaccination was an important predictor of influenza vaccination [52]. There may be several reasons for this. First, people who have already been successfully vaccinated are generally more willing to accept new vaccines. They are certainly more aware of the benefits of vaccination in general and have a better understanding of the procedure and possible side effects. In addition, those who have already been vaccinated with other vaccines may have developed bonds of trust with healthcare professionals, which may also influence their decision to be vaccinated against COVID-19. Finally, people who have received other vaccines as adults are likely more aware of the importance of vaccination in preventing infectious diseases.

The results of this study highlight the strong correlation between confidence in the vaccine and actual uptake. People who have confidence in the vaccine are more likely to be vaccinated. Similar results have been obtained in a study including 23 European Union member states [53] as well as a number of African countries such as Uganda [54], Malawi [27] and the Democratic Republic of Congo [28]. Confidence is a fundamental factor in public acceptance of public health interventions [55]. It is therefore essential to implement strategies to improve confidence in the vaccine to encourage wider actual vaccination against COVID-19 in Benin. These strategies may include specific awareness-raising initiatives led by the country’s most respected health authorities, health professionals, opinion leaders, community, religious or traditional leaders.

Our study showed that location was the third most important factor in determining vaccination status. Compared to the south, people living in the centre are more likely to be vaccinated. It is well known that the refusal of vaccines varies according to the context and the population [56]. The south is the most urbanised region of Benin, where information and communication technologies are the most developed. However, the COVID-19 pandemic was characterised by a high level of infodemia [57,58]. Access to the Internet and social media is more widespread in this area of Benin, but unfortunately these technologies can contribute to vaccine hesitancy because of the speed at which information spreads, the difficulty of verifying the accuracy of online material, and the lack of efforts to combat misinformation. Even a brief exposure to anti-vaccine content can raise perceptions of the risks associated with vaccination and decrease intention to become vaccinated [59,60]. The inhabitants of the southern region were certainly more exposed to the rumours and negative information surrounding the pandemic through social media than people living in other areas. There may also be issues of local leadership in these regions, political relationships between government and local leaders and citizens. Nevertheless, future research must allow us to look more closely at the factors underlying regional disparities and their mechanisms.

We have noted that people who have not received negative information about the vaccine are more likely to be vaccinated. A recent rapid review showed that misinformation on social networks had a negative influence on uptake of the COVID-19 vaccine [61]. A study carried out in Uganda showed similar results [54]. Social media are vectors for mass of information, both truthful and untruthful. In Benin, the pandemic management was severely hampered by notorious misinformation, especially about government measures [57,58,62]. This situation highlights the importance of better controlled communication and education about the vaccine, as well as the importance of combating misinformation and false beliefs likely to harm vaccine uptake.

How individuals perceive waiting times at vaccination centres also affect their decision to be vaccinated. If people think that the waiting time is short and that the process is quick and efficient, they are more likely to be vaccinated. On the other hand, if queues are long and the process tedious, some people may be reluctant to be vaccinated. Similar results have been reported in Ghana [63]. Therefore, health authorities and vaccination centre organisers need to consider this dimension to reduce waiting times and make the vaccination experience as smooth as possible.

Vaccination is often less frequent in people aged 60 and over than in younger people, such as those under 25. This is in line with research carried out in Bangladesh [64], but other studies have found contradictory results [17,18,65]. Older people may be more concerned about the potential risks associated with vaccines because of their frail health. Ease of access to vaccination centres may also be an obstacle for the elderly. It may be difficult for some elderly people to travel or to access vaccination centres.

The results of our study show that risk perception is an important factor in the decision to be vaccinated against COVID-19 in Benin. Indeed, people who think they are more likely to contract the virus are more likely to be vaccinated than those who think they are less likely. This finding is consistent with previous studies [66]. A longitudinal study in South Africa also showed that vaccination was lower among those who thought the vaccine was unsafe [67]. This observation raises crucial questions about the impact of communication about the risks associated with COVID-19 on vaccination behaviour. It underlines the importance of providing clear and precise information about the dangers associated with the disease and the benefits of vaccination, to promote more general adherence to vaccination.

Finally, the results of this study show that the perceived benefits of vaccination play an important role in the decision to be vaccinated. People who see many benefits in vaccination, such as protection against disease, personal safety and contribution to public health, are more likely to be vaccinated. It is therefore essential that awareness and communication campaigns highlight the benefits of vaccination. Similar findings have been reported elsewhere [27,63,65]. Highlighting the direct benefits for individual health, such as reducing the risk of contracting a severe form of the disease, and the benefits for the community, such as contributing to herd immunity and the fight against the spread of the virus, can have a positive impact on public adoption of the vaccine. This has been shown in a mixed-methods study in Niger [68] and a qualitative study in Nigeria [69]. One of the most important factors limiting vaccination is the lack of perceived usefulness of the vaccine, caused by the feeling of not being at risk of the disease. In Benin, traditional healers and charlatans have spread false information, claiming that COVID-19 is simply an aggravated chronic malaria, and claiming that it can be treated with infusions of plants mixed with alcohol [62].

## Limitations

This study has a number of limitations that need to be mentioned. Firstly, as this was a telephone survey, it was not possible to verify the accuracy of all the information provided by the participants, particularly with regard to their vaccination status. We cannot ignore the hypothesis that people eventually over-reported their vaccination status because of social desirability. It should also be noted that we used a cross-sectional approach to analyse the data. This does not allow us to identify temporal relationships of cause and effect. Finally, only people with mobile phones were surveyed, excluding the most marginalised populations.

## Conclusion

The aim of this study was to identify the factors that determine uptake of the first dose of COVID-19 vaccine in Benin. The results highlight the need to consider demographic factors such as geographic location and age, vaccine-related factors such as confidence, perceived risk, perceived benefit and exposure to rumours, and organisational factors such as queuing at vaccination sites when designing vaccination strategies. The results of this study provide valuable information for stakeholders involved in the fight against COVID-19. This information will be of great use in the development of policies and programmes aimed at increasing the effective vaccination coverage of the Beninese population against COVID-19 and other similar pandemics in the future.

## Supporting information

Supplementary material Questionnaire 1

## Data Availability

All data produced in the present study are available upon reasonable request to the authors

## CRediT authorship contribution statement

**Elias Martinien Avahoundje:** Writing–original draft, Writing - Review & Editing, Validation, Software, Investigation, Formal analysis, Data curation, Visualization Methodology. **Christelle Boyi Hounsou:** Writing–original draft, Writing - Review & Editing, Supervision, Validation. **Kéfilath Bello:** Writing–original draft, Writing - Review & Editing, Validation. **Armelle Akouavi Vigan:** Writing–original draft, Writing - Review & Editing. **Christian M Agossou:** Writing - Review & Editing, Investigation. **Mena K. Agbodjavou:** Writing - Review & Editing. **Ibrahima Gaye:** Writing - Review & Editing, Methodology. **Mouhamadou Faly Ba:** Writing - Review & Editing. **Adama Faye:** Writing - Review & Editing, Supervision. **Valéry Ridde:** Writing - Review & Editing, Supervision, Project administration, Funding acquisition, Validation, Methodology, Conceptualization. **Jean Paul Dossou Dossou:** Writing–original draft, Writing - Review & Editing, Supervision, Project administration, Funding acquisition, Validation, Methodology, Conceptualization.

## Acknowledgements

We warmly thank the respondents who agreed to participate in this study by providing valuable information. We would also like to thank the interviewers whose hard work enabled us to collect high quality, usable data. Finally, we would like to thank the entire IT team whose technical support ensured the installation and smooth running of the computer complex used for telephone data collection.

## Funding

This work was supported by the French Development Agency funded as part of the “COVID-19 - Health in common” initiative. It is carried out under the framework of the ARIACOV programme, which stands for “Support to the African Response to the COVID-19 epidemic”

## Appendices

**Table A.1:** Indicator of goodness of fit after CFA and reliability test.

| Variables | Nbr of items | Cronbach's alpha | CFI | RMSEA | SRMR |
| --- | --- | --- | --- | --- | --- |
| Attitude | 5 | 0.7545 | 0.997 | 0.04 | 0.015 |
| Subjective norms | 3 | 0.7852 | 1.000 | 0.00 | 0.000 |
| Behavioral control<br>(autonomy) | 1 | - | - | - | - |
| Behavioral control<br>(capacity) | 4 | 0.5671 | 0.983 | 0.05 | 0.023 |
| Perceived effectiveness | 2 | 0.6599 | 1.000 | 0.00 | 0.000 |
| Perceived benefits | 3 | 0.9085 | 1.000 | 0.00 | 0.000 |
| Perceived risks | 4 | 0.6294 | 0.998 | 0.03 | 0.012 |
| Negative emotions | 3 | 0.9110 | 1.000 | 0.00 | 0.000 |
| Positive emotions | 4 | 0.6577 | 1.000 | 0.013 | 0.012 |
| Vaccine confidence | 1 | - | - | - | - |
| Provider trust | 1 | - | - | - | - |

## Notes

### Competing Interest Statement

The authors have declared no competing interest.

### Author Declarations

This study was approved by the Local Ethics Committee for Biomedical Research of the University of Parakou in Benin under number: 0308/CLERB-UP/P/SP/R/SA.

## References

1. World Health Organisation (WHO). WHO Coronavirus Disease (COVID-19) dashboard with vaccination Data. [Internet]. 2024 [cited 2024 Feb 28]. Available from: https://data.who.int/dashboards/covid19/deaths?n=c

2. Barrett ADT, Titball RW, MacAry PA, Rupp RE, Messling V von, Walker DH, et al. The rapid progress in COVID vaccine development and implementation. npj Vaccines. 2022 Feb;7(1):20.

3. Fiolet T, Kherabi Y, MacDonald CJ, Ghosn J, Peiffer-Smadja N. Comparing COVID-19 vaccines for their characteristics, efficacy and effectiveness against SARS-CoV-2 and variants of concern: a narrative review. Clinical Microbiology and Infection. 2022 Feb;28(2):202–21.

4. Tenforde MW, Self WH, Adams K, Gaglani M, Ginde AA, McNeal T, et al. Association Between mRNA Vaccination and COVID-19 Hospitalization and Disease Severity. JAMA. 2021 Nov;326(20):2043.

5. Lazarus JV, Wyka K, White TM, Picchio CA, Gostin LO, Larson HJ, et al. A survey of COVID-19 vaccine acceptance across 23 countries in 2022. Nature Medicine. 2023 Feb;29(2):366–75.

6. Samba E, Nkrumah F, Leke R. Getting Polio Eradication Back on Track in Nigeria. New England Journal of Medicine. 2004 Feb;350(7):645–6.

7. Katz I, Glandon D, Wong W, Kargbo B, Ombam R, Singh S, et al. Lessons learned from stakeholder-driven sustainability analysis of six national {HIV} programmes. Health policy and planning. 2014 May;29(3):379–87.

8. Kim JH, Marks F, Clemens JD. Looking beyond COVID-19 vaccine phase 3 trials. Nature Medicine. 2021 Feb;27(2):205–11.

9. Omar DI, Hani BM. Attitudes and intentions towards COVID-19 vaccines and associated factors among Egyptian adults. Journal of Infection and Public Health [Internet]. 2021 Oct;14(10):1481–8. Available from: 10.1016/j.jiph.2021.06.019 https://linkinghub.elsevier.com/retrieve/pii/S1876034121001854

10. Avahoundje EM, Dossou JP, Vigan A, X Igv, undefined 2022. Factors associated with COVID-19 vaccine intention in Benin in 2021: A cross-sectional study. Elsevier [Internet]. Available from: https://www.sciencedirect.com/science/article/pii/S2590136222000973

11. Yan E, Lai DWL, Ng HKL, Lee VWP. Predictors of COVID-19 actual vaccine uptake in Hong Kong: A longitudinal population-based survey. SSM Popul Health [Internet]. 2022;18:101130. Available from: 10.1016/j.ssmph.2022.101130 https://www.ncbi.nlm.nih.gov/pubmed/35620485 https://www.ncbi.nlm.nih.gov/pmc/articles/PMC9119715

12. Wouters OJ, Shadlen KC, Salcher-Konrad M, Pollard AJ, Larson HJ, Teerawattananon Y, et al. Challenges in ensuring global access to COVID-19 vaccines: production, affordability, allocation, and deployment. The Lancet. 2021 Mar;397(10278):1023–34.

13. Markovic-Denic L, Nikolic V, Pavlovic N, Maric G, Jovanovic A, Nikolic A, et al. Changes in Attitudes toward COVID-19 Vaccination and Vaccine Uptake during Pandemic. Vaccines. 2023 Jan;11(1):147.

14. Pedersen B, Thanel K, Kouakou A, Zo J, Ouattara M, Gbeke D, et al. Identifying Drivers of COVID-19 Vaccine Uptake among Residents of Yopougon Est, Abidjan, Côte d’Ivoire. Vaccines [Internet]. 2022 Dec;10(12):2101. Available from: https://www.mdpi.com/2076-393X/10/12/2101

15. Corcoran KE, Scheitle CP, DiGregorio BD. Christian nationalism and COVID-19 vaccine hesitancy and uptake. Vaccine [Internet]. 2021;39(45):6614–21. Available from: 10.1016/j.vaccine.2021.09.074 https://www.ncbi.nlm.nih.gov/pubmed/34629205 https://www.ncbi.nlm.nih.gov/pmc/articles/PMC8489517

16. Galanis P, Vraka I, Siskou O, Konstantakopoulou O, Katsiroumpa A, Kaitelidou D. Uptake of COVID-19 Vaccines among Pregnant Women: A Systematic Review and Meta-Analysis. Vaccines [Internet]. 2022 May;10(5):766. Available from: https://www.mdpi.com/2076-393X/10/5/766

17. Abebe EC, Tiruneh GA, Adela GA, Ayele TM, Muche ZT, T/Mariam AB, et al. COVID-19 vaccine uptake and associated factors among pregnant women attending antenatal care in Debre Tabor public health institutions: A cross-sectional study. Frontiers in Public Health [Internet]. 2022 Jul;10:919494. Available from: https://www.frontiersin.org/articles/10.3389/fpubh.2022.919494/full

18. Terefa DR, Shama AT, Feyisa BR, Desisa AE, Geta ET, Cheme MC, et al. COVID-19 Vaccine Uptake and Associated Factors Among Health Professionals in Ethiopia. Infect Drug Resist [Internet]. 2021;14:5531–41. Available from: 10.2147/IDR.S344647 https://www.ncbi.nlm.nih.gov/pubmed/34984008 https://www.ncbi.nlm.nih.gov/pmc/articles/PMC8702775

19. Iversen J, Wand H, Kemp R, Bevan J, Briggs M, Patten K, et al. Uptake of COVID-19 vaccination among people who inject drugs. Harm Reduction Journal [Internet]. 2022 Dec;19(1):59. Available from: https://harmreductionjournal.biomedcentral.com/articles/10.1186/s12954-022-00643-3

20. Anino CO, Wandera I, Masimba ZO, Kirui CK, Makero CS, Omari PK, et al. Determinants of Covid-19 vaccine uptake among the elderly aged 58 years and above in Kericho County, Kenya: Institution based cross sectional survey. Oliveira EF de, editor. PLOS Global Public Health [Internet]. 2023 Sep;3(9):e0001562. Available from: https://dx.plos.org/10.1371/journal.pgph.0001562

21. Memedovich A, Orr T, Hollis A, Salmon C, Hu J, Zinszer K, et al. Social network risk factors and COVID-19 vaccination: A cross-sectional survey study. Vaccine [Internet]. 2024 Feb;42(4):891–911. Available from: https://linkinghub.elsevier.com/retrieve/pii/S0264410×24000057

22. Alya WA, Maraqa B, Nazzal Z, Odeh M, Makhalfa R, Nassif A, et al. COVID-19 vaccine uptake and its associated factors among Palestinian healthcare workers: Expectations beaten by reality. Vaccine [Internet]. 2022 Jun;40(26):3713–9. Available from: https://linkinghub.elsevier.com/retrieve/pii/S0264410×22006107

23. Shaw J, Stewart T, Anderson KB, Hanley S, Thomas SJ, Salmon DA, et al. Assessment of US Healthcare Personnel Attitudes Towards Coronavirus Disease 2019 (COVID-19) Vaccination in a Large University Healthcare System. Clinical Infectious Diseases. 2021 Nov;73(10):1776–83.

24. Galanis P, Vraka I, Katsiroumpa A, Siskou O, Konstantakopoulou O, Katsoulas T, et al. COVID-19 Vaccine Uptake among Healthcare Workers: A Systematic Review and Meta-Analysis. Vaccines (Basel) [Internet]. 2022;10(10). Available from: 10.3390/vaccines10101637 https://www.ncbi.nlm.nih.gov/pubmed/36298502 https://www.ncbi.nlm.nih.gov/pmc/articles/PMC9610263

25. Najjar M, Albuaini S, Fadel M, Mohsen F, Najjar G, Assaf D, et al. Covid-19 vaccination reported side effects and hesitancy among the Syrian population: a cross-sectional study. Annals of Medicine [Internet]. 2023 Dec;55(2). Available from: https://www.tandfonline.com/doi/full/10.1080/07853890.2023.2241351

26. Galanis P, Vraka I, Katsiroumpa A, Siskou O, Konstantakopoulou O, Zogaki E, et al. Psychosocial Predictors of COVID-19 Vaccine Uptake among Pregnant Women: A Cross-Sectional Study in Greece. Vaccines [Internet]. 2023 Jan;11(2):269. Available from: https://www.mdpi.com/2076-393X/11/2/269

27. Madhlopa QK, Mtumbuka M, Kumwenda J, Illingworth TA, Van Hout MC, Mfutso-Bengo J, et al. Factors affecting COVID-19 vaccine uptake in populations with higher education: insights from a cross-sectional study among university students in Malawi. BMC Infect Dis. 2024 Aug 21;24(1):848.

28. Akilimali PZ, Egbende L, Kayembe DM, Kabasubabo F, Kazenza B, Botomba S, et al. COVID-19 Vaccine Coverage and Factors Associated with Vaccine Hesitancy: A Cross-Sectional Survey in the City of Kinshasa, Democratic Republic of Congo. Vaccines. 2024 Feb 12;12(2):188.

29. Hao F. A cross-national study of multilevel determinants on public fully vaccination against COVID-19. Health & Place [Internet]. 2023 Jan;79:102963. Available from: https://linkinghub.elsevier.com/retrieve/pii/S1353829222002246

30. Altulaihi BA, Alharbi KG, Alaboodi TA, Alkanhal HM, Alobaid MM, Aldraimly MA. Factors and Determinants for Uptake of COVID-19 Vaccine in a Medical University in Riyadh, Saudi Arabia. Cureus [Internet]. 2021 Sep;13(9):e17768. Available from: https://www.cureus.com/articles/68797-factors-and-determinants-for-uptake-of-covid-19-vaccine-in-a-medical-university-in-riyadh-saudi-arabia

31. Gouvernement de la République du Bénin. Campagne accélérée de vaccination contre la Covid-19 au Béninl: Les leaders d’opinion sensibilisés [Internet]. 2022. Available from: https://www.gouv.bj/article/1692/campagne-acceleree-vaccination-contre-covid-19-benin-leaders-opinion-sensibilises/

32. Vignigbé M, Maccaro A, Rifà PC, Piaggio D, Pecchia L. Précaution ou prévention? Des responsabilités de la politique et de la société pendant la gestion de la pandémie de COVID-19 au Bénin. Canadian Journal of Development Studies / Revue canadienne d’études du développement. 2024 Jan;1–16.

33. Kislaya I, Andrianarimanana DK, Marchese V, Hosay L, Rivomalala R, Holinirina R, et al. Drivers of COVID-19 vaccine uptake among rural populations in Madagascar: a cross-sectional study. BMC Public Health. 2024 Oct 17;24(1):2868.

34. Daniel DA, Akwaras NA, De-Kaa NLP, Nwaeze C, Swende LT, Ornguga BO. Predictors of COVID-19 Vaccine Uptake in a Tertiary Hospital Community, North Central, Nigeria. West Afr J Med. 2024 May 31;41(5):542–7.

35. Kebede SD, Aytenew TM. Attitude, knowledge, and predictors of COVID-19 vaccine uptake among health care providers in South Gondar public hospitals, North Central Ethiopia: multi-facility based study. Pan Afr Med J. 2022;41:194.

36. Padonou SGR, Glèlè CK, Vaccines MA, undefined 2023. Assessment of COVID-19 Vaccine Acceptance and Its Associated Factors during the Crisis: A Community-Based Cross-Sectional Study in Benin. mdpi.com [Internet]. Available from: https://www.mdpi.com/2076-393X/11/6/1104

37. Institut National de la Statistique et de la Démographie (INStaD). Projections démographiques de 2014 à 2063 et perspectives de la demande sociale de 2014 à 2030 au Bénin. INStaD; 2022.

38. United Nations Development Programme. Human Development Report 2021/2022: Uncertain times, unsettled lives.Shaping our future in a transforming world. UNDP; 2022.

39. Institut National de la Statistique et de la Démographie (INStaD). Enquête par Grappes à Indicateurs Multiples, Bénin, 2021-2022, Rapport des résultats de l’enquête. INStaD; 2023.

40. OMS. Le Bénin reçoit ses premières doses de vaccins contre la COVID-19 via le mécanisme COVAX [Internet]. 2021. Available from: https://www.afro.who.int/fr/countries/benin/news/le-benin-recoit-ses-premieres-doses-de-vaccins-contre-la-covid-19-le-mecanisme-covax

41. Gaye I, Ridde V, Avahoundjea EM, Ba MF, Dossou JP, Diallo AI, et al. Cross-sectional study on intention to be vaccinated against Coronavirus Disease 2019 (COVID-19) in Benin and Senegal: A structural equation modeling (SEM). Asweto CO, editor. PLOS Glob Public Health [Internet]. 2024 Mar 18 [cited 2024 Aug 30];4(3):e0002868. Available from: https://dx.plos.org/10.1371/journal.pgph.0002868

42. Deville JC. Une théorie des enquêtes par quotas. Techniques d’enquête, Statistique Canada [Internet]. 1991;12(2):177–95. Available from: https://www150.statcan.gc.ca/n1/fr/pub/12-001-x/1991002/article/14504-fra.pdf?st=a165SGXA

43. U. S. Census Bureau. Benin Annual Five-Year Age Group Population Estimates by Sex for 2015 to 2030: National, and First- and Second-Order Administrative Divisions [Internet]. Available from: https://www2.census.gov/programs-surveys/international-programs/tables/time-series/pepfar/benin.xlsx

44. Norman P, Conner M. Health Behavior Ill. In: Reference Module in Neuroscience and Biobehavioral Psychology [Internet]. Elsevier; 2017. p. 1–37. Available from: https://linkinghub.elsevier.com/retrieve/pii/B9780128093245051439

45. Green EC, Murphy EM, Gryboski K. The Health Belief Model. The Wiley Encyclopedia of Health Psychology [Internet]. 2021 Sep;2(1):211–4. Available from: https://onlinelibrary.wiley.com/doi/10.1002/9781119057840.ch68

46. Ursachi G, Horodnic IA, Zait A. How Reliable are Measurement Scales? External Factors with Indirect Influence on Reliability Estimators. Procedia Economics and Finance. 2015;20:679–86.

47. Acock AC. Discovering Structural Equation Modeling Using Stata. Stata Press books [Internet]. 2013; Available from: https://ideas.repec.org/b/tsj/spbook/dsemus.html

48. Schermelleh-Engel K, Moosbrugger H, Müller H. Evaluating the Fit of Structural Equation Models: Tests of Significance and Descriptive Goodness-of-Fit Measures. Methods of Psychological Research [Internet]. 2003;8(2):23–74. Available from: https://psycharchives.org/en/item/1a8dea48-0285-4dac-a612-9dc0ff2532f6

49. Midi H, Sarkar SK, Rana S. Collinearity diagnostics of binary logistic regression model. Journal of Interdisciplinary Mathematics [Internet]. 2010 Jun;13(3):253–67. Available from: http://www.tandfonline.com/doi/abs/10.1080/09720502.2010.10700699

50. Lomeli A, Escoto AA, Reyes B, Burola MLM, Tinoco-Calvillo S, Villegas I, et al. Factors associated with COVID-19 vaccine uptake in a US/Mexico border community: demographics, previous influenza vaccination, and trusted sources of health information. Front Public Health [Internet]. 2023 Jul 27 [cited 2024 Oct 3];11:1163617. Available from: https://www.frontiersin.org/articles/10.3389/fpubh.2023.1163617/full

51. Han K, Hou Z, Tu S, Liu M, Chantler T, Larson H. Factors Influencing Childhood Influenza Vaccination: A Systematic Review. Vaccines [Internet]. 2024 Feb 23 [cited 2024 Oct 3];12(3):233. Available from: https://www.mdpi.com/2076-393X/12/3/233

52. Sibanda M, Burnett RJ, Godman B, Meyer JC. Vaccine uptake, associated factors and reasons for vaccination status among the South African elderly; findings and next steps. Carels V, editor. PLoS ONE. 2024 Dec 4;19(12):e0314098.

53. Cartanyà-Hueso À, Martínez-Sánchez JM, Martín-Sánchez JC, Lidón-Moyano C, Pérez-Martín H, González-Marrón A. Relationship between double COVID-19 vaccine uptake and trust in effectiveness and safety of vaccination in general in 23 Member states of the European Union: an ecological study. Vaccine. 2022 Jul;40(32):4334–8.

54. Kyakuwa N, Abaasa A, Mpooya S, Kalutte H, Atuhairwe C, Perez L, et al. Non-uptake of COVID-19 vaccines and reasons for non-uptake among healthcare workers in Uganda: a cross-sectional study. BMC Health Services Research [Internet]. 2024 May;24(1):663. Available from: https://bmchealthservres.biomedcentral.com/articles/10.1186/s12913-024-11137-2

55. Gilson L. Trust and the development of health care as a social institution. Social Science & Medicine. 2003 Apr;56(7):1453–68.

56. MacDonald NE. Vaccine hesitancy: Definition, scope and determinants. Vaccine [Internet]. 2015 Aug;33(34):4161–4. Available from: https://linkinghub.elsevier.com/retrieve/pii/S0264410×15005009

57. Houngnihin RA, Ridde V, Traverson L, Zinszer K. Le port obligatoire du masque dans la réponse à la pandémie de Covid-19 au Bénin. Entre popularité et scepticisme in Hôpitaux et santé publique face à la pandémie de Covid-19 [Internet]. QuébecLJ: Éditions science et bien commun; [cited 2024 Dec 18]. Available from: https://scienceetbiencommun.pressbooks.pub/hospicovid/

58. Akpi E, Agbodjavou M, Bedie V, Kougblenou O, Mensah Coffi Mahugnon André A, Kpatchavi A, et al. Rumeurs, attitudes et pratiques sur la vaccination dans un contexte de maladie dite «LJimportéeLJ». Exemple du covid-19 en milieu Adja-Fon au sud-ouest du Bénin. Antropo. 2023 Dec;46:67–82.

59. Stewart R, Madonsela A, Tshabalala N L, Theunissen N. The importance of social media users’ responses in tackling digital COVID-19 misinformation in Africa. DIGITAL HEALTH [Internet]. 2022 Jan [cited 2024 Oct 3];8:205520762210850. Available from: http://journals.sagepub.com/doi/10.1177/20552076221085070

60. Betsch C, Renkewitz F, Betsch T, Ulshöfer C. The Influence of Vaccine-critical Websites on Perceiving Vaccination Risks. J Health Psychol [Internet]. 2010 Apr [cited 2024 Oct 3];15(3):446–55. Available from: https://journals.sagepub.com/doi/10.1177/1359105309353647

61. Skafle I, Nordahl-Hansen A, Quintana DS, Wynn R, Gabarron E. Misinformation About COVID-19 Vaccines on Social Media: Rapid Review. Journal of Medical Internet Research. 2022 Aug;24(8):e37367.

62. MFWA. Les Médias et les Fake News sur la COVID-19LJ: Cas du Benin [Internet]. Media Foundation For West Africa; 2020. Available from: https://www.mfwa.org/wp-content/uploads/2020/12/Rapport-Final-Medias-et-Covid-19-Benin-rvd-e-1.pdf

63. Amoah JO, Abraham SA, Adongo CA, Sekimpi DK, Adukpo DC, Obiri-Yeboah D, et al. Determinants of COVID-19 vaccine uptake: evidence from a vulnerable global South setting. BMC Research Notes [Internet]. 2024 Mar;17(1):94. Available from: https://bmcresnotes.biomedcentral.com/articles/10.1186/s13104-024-06736-5

64. Abedin M, Islam MA, Rahman FN, Reza HM, Hossain MZ, Hossain MA, et al. Willingness to vaccinate against COVID-19 among Bangladeshi adults: Understanding the strategies to optimize vaccination coverage. Kabir E, editor. PLOS ONE [Internet]. 2021 Apr;16(4):e0250495. Available from: https://dx.plos.org/10.1371/journal.pone.0250495

65. Pijpers J, Roon A van, Roekel C van, Labuschagne L, Smagge B, Ferreira JA, et al. Determinants of COVID-19 Vaccine Uptake in The Netherlands: A Nationwide Registry-Based Study. Vaccines [Internet]. 2023 Aug;11(9):1409. Available from: https://www.mdpi.com/2076-393X/11/9/1409

66. Viswanath K, Bekalu M, Dhawan D, Pinnamaneni R, Lang J, McLoud R. Individual and social determinants of COVID-19 vaccine uptake. BMC Public Health. 2021 Dec;21(1):818.

67. Maughan-Brown B, Eyal KC, Njozela L, Buttenheim AM. Predictors of COVID-19 vaccine uptake among adults in South Africa: multimethod evidence from a population-based longitudinal study. BMJ Global Health [Internet]. 2023 Aug;8(8):e012433. Available from: https://gh.bmj.com/lookup/doi/10.1136/bmjgh-2023-012433

68. Seytre B, Chaibou S, Simon B. The Drivers of Low Vaccination Utilization in Niger. The American Journal of Tropical Medicine and Hygiene. 2024 Mar;110(3):529–33.

69. Wonodi C, Obi-Jeff C, Adewumi F, Keluo-Udeke SC, Gur-Arie R, Krubiner C, et al. Conspiracy theories and misinformation about COVID-19 in Nigeria: Implications for vaccine demand generation communications. Vaccine. 2022 Mar;40(13):2114–21.

