## Supplementary material Questionnaire 1 for "Predictors of first-dose COVID-19 vaccine uptake in Benin: Evidence from a cross-sectional study based on a telephonic survey"

### Supplementary material 1

#### CAP-CoV-Africa - General population questionnaire – Benin.

**Ethics :** Introductory research question and acceptance to answer and be contacted further later for more in-depth discussions

**Date and time of collection:**

____________________________________________

**Collection agent code: _ _ _**

***Investigator instructions:*** *Unless otherwise specified, list all possible answers (to except for the modalities Don't Know or NSP and Refusal).*

**SECTION: Consent and general characteristics**

| label | terms and conditions | Variables code |
|---|---|---|
| **Q.1 Would you have some minutes hasagree to answer my questions?** | 1. Yes<br>2. Refusal<br>3. Impossibilitylinguistic (does not speak any of the languages spoken by investigators)<br>4. Impossibilitymental (the person is not able to consent)<br>*If different from Yes, stop the questionnaire.* | **Q1** |

*Note: Questions Q2 to Q8 are asked only of new respondents who did not take part in phase 1.*

| VARIABLES | TERMS AND CONDITIONS | Code |
|---|---|---|
| Q2 Department | 1. Alibori<br>2. Atacora<br>3. Atlantic<br>4. Borgou<br>5. Hills<br>6. Couffo<br>7. Donga<br>8. Coastline<br>9. Mono<br>10. Oueme<br>11. Plateau<br>12. Zou | department |
| Q3 Age (in years) | | age |
| Q4 Sex | 1. Male<br>2. Female | gender |

| | | |
|---|---|---|
| Q5 Marital status | 1. Married/monogamous<br>2. Married/polygamous<br>3. Cohabiting as a couple without being married<br>4. Divorced<br>5. Widowed<br>6. Single | Sit_matri |
| Q6 Level of education | 1. Uneducated<br>2. Primary<br>3. Secondary<br>4. University | Level_instruction |
| Q7 Financially, would you say? that ? | 1. You are very comfortable<br>2. Your income is sufficient<br>3. You are poor<br>4. You are very poor | Financial_level |

Q8 For each of the following goods, can you tell me if your household is equipped with one?

| | | |
|---|---|---|
| Radio | 1. Yes<br>2. No<br>99. NSP | Radio |
| Television | 1. Yes<br>2. No<br>99. NSP | television |
| Fridge | 1. Yes<br>2. No<br>99. NSP | fridge |
| Running water (tap) | 1. Yes in accommodation with individual meter<br>2. Yes shared, public or without individual meter<br>3. No<br>99. NSP | water |
| Electricity | 1. Yes with meterindividual, bandgenerator or panelsolar<br>2. Yes without meter individual<br>3. No<br>99. NSP | Electro |

| **SECTION: Vaccination** |
|---|

**We will now ask you questions about the available COVID-19 vaccines.**

| | **Possible choices** | **Codification** |
|---|---|---|
| **Q9 Do you know that there are vaccines against COVID-19?** | 1. Yes<br>2. No<br>99. I don't know | **Q9** |
| **Q10 Have you received any vaccinations since you were over 18 years old?** | 1. Yes<br>2. No<br>99. I don't know | **Q10** |
| **Q11 Have you received a COVID-19 vaccine?** | 1. Yes<br>2. No<br>99. I prefer not to answer | **Q11** |
| **Q12 If yes, what vaccine did you receive?** | 1. Johnson&Johnson (Janssen)<br>2. Sinovac (CoronaVac)<br>3. AstraZeneca (Covishield or Vaxzevria)<br>4. Pfizer<br>5. Moderna (Spikevax),<br>6. Sputnik V<br>99. I don't know<br>If answer 1 (Johnson&Johnson), skip the next question | **Q12** |
| **Q13 Have you already received all doses of this vaccine?** | 1. Yes<br>2. No<br>99. I don't know | **Q13** |
| **Q14 Have you had any effects secondary effects related to one of the doses of vaccine ?** | 1. Yes<br>2. No<br>99. I don't know | **Q14** |

| | | |
|---|---|---|
| Q15 How do you rate the severity of the side effects received? | 1. Very serious<br>2. Severe<br>3. Less serious<br>4. Not at all serious<br>99. You don't know | Q15 |

| **Q.16 ATTITUDE TOWARDS THE VACCINE** | | |
|---|---|---|
| a. I think it is important to get vaccinated | 5. Totally agree<br>4. Okay<br>3. Neither agree nor disagreement<br>2. Disagree<br>1. Strongly disagree | **Q16a** |
| b. I think it is useful to get vaccinated to protect yourself against COVID-19 | 5. Totally agree<br>4. Okay<br>3. Neither agree nor disagreement<br>2. Disagree<br>1. Strongly disagree | **Q16b** |
| c. I think it is responsible to get vaccinated against COVID-19 | 5. Totally agree<br>4. Okay<br>3. Neither agree nor disagreement<br>2. Disagree<br>1. Strongly disagree | **Q16c** |
| d. I believe that the COVID-19 vaccine does not pose a health risk | 5. Totally agree<br>4. Okay<br>3. Neither agree nor disagreement<br>2. Disagree<br>1. Strongly disagree | **Q16d** |
| e. I think it is advisable to get vaccinated against COVID-19 | 5. Totally agree<br>4. Okay<br>3. Neither agree nor disagreement<br>2. Disagree<br>1. Strongly disagree | **Q16e** |
| **Q.17 INTENTIONS** | | |

| | | |
|---|---|---|
| a. I intend to get vaccinated against COVID-19 | 5. Totally agree<br>4. Okay<br>3. Neither agree nor disagreement<br>2. Disagree<br>1. Strongly disagree | **Q17a** |
| b. What do you **would motivate**to get vaccinated? | Open answer: ………………… | **Q17b** |
| c. I plan to recommend the coronavirus vaccine to those around me | 5. Totally agree<br>4. Okay<br>3. Neither agree nor disagreement<br>2. Disagree<br>1. Strongly disagree | **Q17c** |
| **Q.18 PERCEIVED BENEFIT** | | |
| a. Getting vaccinated against COVID-19 will help protect me from the virus | 5. Totally agree<br>4. Okay<br>3. Neither agree nor disagreement<br>2. Disagree<br>1. Strongly disagree | **Q18a** |
| b. Getting vaccinated will help fight the spread of coronavirus | 5. Totally agree<br>4. Okay<br>3. Neither agree nor disagreement<br>2. Disagree<br>1. Strongly disagree | **Q18b** |
| c. Getting vaccinated against COVID-19 will help protect my loved ones from the virus | 5. Totally agree<br>4. Okay<br>3. Neither agree nor disagreement<br>2. Disagree<br>1. Strongly disagree | **Q18c** |
| **Q.19 PERCEIVED RISK** | | |
| a. I believe that those who created the COVID-19 vaccine will ensure its safety. | 5. Totally agree<br>4. Okay<br>3. Neither agree nor disagreement<br>2. Disagree<br>1. Strongly disagree | **Q19a** |

| | | |
|---|---|---|
| b. The coronavirus vaccine could put my health at risk | 5. Totally agree<br>4. Okay<br>3. Neither agree nor disagreement<br>2. Disagree<br>1. Strongly disagree | **Q19b** |
| c. THE vaccine against THE Coronavirus could have side effects | 5. Totally agree<br>4. Okay<br>3. Neither agree nor disagreement<br>2. Disagree<br>1. Strongly disagree | **Q19c** |
| d. Vaccination in children aged 12 to 18 is dangerous for their health | 5. Totally agree<br>4. Okay<br>3. Neither agree nor disagreement<br>2. Disagree<br>1. Strongly disagree | **Q19d** |
| **Q.20 PERCEIVED EFFECTIVENESS** | | |
| a. I think that if I get vaccinated against COVID-19, it is unlikely that I will become infected | 5. Totally agree<br>4. Okay<br>3. Neither agree nor disagreement<br>2. Disagree<br>1. Strongly disagree | **Q20a** |
| b. I think the vaccine will reduce the risk of getting COVID-19 | 5. Totally agree<br>4. Okay<br>3. Neither agree nor disagreement<br>2. Disagree<br>1. Strongly disagree | **Q20b** |
| **Q.21 EMOTIONS** | | |
| a. Faced with the coronavirus, I have<br>**fear** | 5. Totally agree<br>4. Okay<br>3. Neither agree nor disagreement<br>2. Disagree<br>1. Strongly disagree | **Q21a** |

| | | |
|---|---|---|
| b. Faced with the coronavirus, I feel anxious | 5. Totally agree<br>4. Okay<br>3. Neither agree nor disagreement<br>2. Disagree<br>1. Strongly disagree | **Q21b** |
| c. Faced with the coronavirus, I feel worried | 5. Totally agree<br>4. Okay<br>3. Neither agree nor disagreement<br>2. Disagree<br>1. Strongly disagree | **Q21c** |
| d. Faced with the coronavirus, I have a lot of hope | 5. Totally agree<br>4. Okay<br>3. Neither agree nor disagreement<br>2. Disagree<br>1. Strongly disagree | **Q21d** |
| e. Faced with the coronavirus, I feel optimistic | 5. Totally agree<br>4. Okay<br>3. Neither agree nor disagreement<br>2. Disagree<br>1. Strongly disagree | **Q21e** |
| f. Faced with the coronavirus, I feel enthusiastic | 5. Totally agree<br>4. Okay<br>3. Neither agree nor disagreement<br>2. Disagree<br>1. Strongly disagree | **Q21f** |
| g. Faced with the coronavirus, I feel confident | 5. Totally agree<br>4. Okay<br>3. Neither agree nor disagreement<br>2. Disagree<br>1. Strongly disagree | **Q21g** |
| **Q.22 SUBJECTIVE NORMS** | | |

| | | |
|---|---|---|
| a. Most important people around me (family, friends) think I should get the COVID-19 vaccine | 5. Totally agree<br>4. Okay<br>3. Neither agree nor disagreement<br>2. Disagree<br>1. Strongly disagree | **Q22a** |
| b. People whose opinions are important to me approve of getting the coronavirus vaccine | 5. Totally agree<br>4. Okay<br>3. Neither agree nor disagreement<br>2. Disagree<br>1. Strongly disagree | **Q22b** |
| c. Nursing staff<br>think i should get the COVID vaccine | 5. Totally agree<br>4. Okay<br>3. Neither agree nor disagreement<br>2. Disagree<br>1. Strongly disagree | **Q22c** |

**BEHAVIOUR TOWARDS VACCINE**

| **Q.23 BEHAVIORAL CONTROL (ability)** | | |
|---|---|---|
| a. It is easy for me to get to the vaccination site to get vaccinated against coronavirus if I want to | 5. Totally agree<br>4. Okay<br>3. Neither disagree nor agree<br>2. Disagree<br>1. Strongly disagree | **Q23a** |
| b. I have the financial means to go to the vaccination site | 5. Totally agree<br>4. Okay<br>3. Neither disagree nor agree<br>2. Disagree<br>1. Strongly disagree | **Q23b** |
| c. I think it is easy for me to get vaccinated against coronavirus if campaigns of vaccination are organized. | 5. Completely confident<br>4. Confident<br>3. Neither agree nor disagree<br>2. not confident<br>1. not at all confident | **Q23c** |

| | | |
|---|---|---|
| d. I feel completely able to find the information I have need on vaccine against coronavirus | 5. Completely confident<br>4. Confident<br>3. Neither agree nor disagree<br>2. not confident<br>1. not at all confident | **Q23d** |
| **Q.23 BEHAVIORAL CONTROL (autonomy)** | | |
| f. It is up to me to decide whether I want to get a coronavirus vaccine | 5. Totally agree<br>4. Okay<br>3. Neither agree nor disagree<br>2. Disagree<br>1. Strongly disagree | **Q23f** |

| | | |
|---|---|---|
| **Q24 CONFIDENCE IN COVID-19 VACCINES** | | |
| a. How much do you trust healthcare providers who administer COVID-19 vaccinations? Would you say that you trust them... | 5. Completely confident<br>4. Confident<br>3. Neither agree nor disagree<br>2. not confident<br>1. not at all confident | **Q24a** |
| Q24b How much do you trust vaccines? against COVID-19? | 5. Completely confident<br>4. Confident<br>3. Neither confident nor not confident<br>2. not confident<br>1. not at all confident | **Q24b** |

| | | |
|---|---|---|
| **Q 25 Organization in vaccination locations** | | |
| a. The length of the queue for vaccination against covid 19 is acceptable | 5. Totally agree<br>4. Okay<br>3. Neither agree nor disagree<br>2. Disagree<br>1. Strongly disagree | **Q25a** |

| | | |
|---|---|---|
| b. THE schedules opening of the centers of vaccinations are adequate For me allow me to get vaccinated | 5. Totally agree<br>4. Okay<br>3. Neither agree nor disagree<br>2. Disagree<br>1. Strongly disagree | **Q25b** |

| Q25 Information on vaccines | | |
|---|---|---|
| a. Over the next few months, I will regularly learn about COVID-19 vaccines. | 5. Totally agree<br>4. Okay<br>3. Neither agree nor disagreement<br>2. Disagree<br>1. Strongly disagree | **Q25a** |
| b. I will seek information about coronavirus vaccines to better understand it. | 5. Totally agree<br>4. Okay<br>3. Neither agree nor disagreement<br>2. Disagree<br>1. Strongly disagree | **Q25b** |
| c. I will read the information I receive about COVID-19 vaccines through social media | 5. Totally agree<br>4. Okay<br>3. Neither agree nor disagreement<br>2. Disagree<br>1. Strongly disagree | **Q25c** |
| d. Have you seen or heard anything bad about the COVID-19 vaccines? | 1. Yes<br>2. No | **Q25d1** |
| | If yes, please specify<br>……………………… | **Q25d2** |

#### SECTION: End of questionnaire and reminder

**Your answers will be very valuable to us in improving public health programs.**

**Thank you for giving me all this time.**
**Thank you. I wish you a great end of the day.**

**If you have any questions about COVID-19, you can call 136 for free**
